# Screening, diagnosis and hospitalization of breast and cervical cancer in Brazil from 2010 to 2022: a time-series study

**DOI:** 10.1101/2022.11.09.22282147

**Authors:** Gustavo Nepomuceno Capistrano, Thiffany Nayara Bento de Morais, Ketyllem Tayanne da Silva Costa, Ana Luiza Santos Quirino, Roberta Letícia Pimentel da Costa, Fábia Barbosa de Andrade

**Author notes:** Corresponding Author (KTSC). These authors contributed equally to this work.

## Abstract

In Brazil, during the pandemic caused by COVID-19, screening for breast and cervical cancers was postponed or interrupted due to the prevailing health conditions. These neoplasms, however, are responsible for high morbidity and mortality among women in Brazil and have a major impact on the quality of life of this population and on public health. Thus, this study aims to evaluate the epidemiological behavior of hospitalization for cervical and breast cancer in Brazilian women, as well as the trend of screening tests and diagnosis of breast and cervical cancer in the years 2010 to 2022 in Brazil. This is an ecological research of time series, based on secondary data obtained from information systems of the country, about hospital admissions for breast and cervical cancer and screening methods used for these tumors. The data were analyzed in the Joinpoint Regression Program, to obtain the linear regression and temporal analysis of the variables. As results, there is a decrease in hospitalization rates for cervical cancer between the years 2010 and 2015 and a subsequent increase in 2019. Regarding breast cancer, there was an increase in hospitalizations, until reaching a peak in 2019. For both, in the pandemic years, between 2020 and 2022, there is a decrease in Brazil and in all its regions. As for the tracking of these diseases, it was observed that the performance of mammograms and preventive tests showed a similar behavior, in which there is a higher supply of these tests until 2019 and a drop during the pandemic period. This leads to the conclusion that even though Brazil has several policies for screening and early diagnosis of these diseases, there is still instability in the offer of tests and that there was a precariousness in this area during the pandemic.

## Introduction

Cancer is the main public health problem in the world and is better observed in developing countries. Factors such as older age, low socioeconomic conditions, and poor lifestyle habits are directly related to its incidence and mortality^1^.

According to estimates, breast cancer is the most common cancer in women in all regions of Brazil, except for non-melanoma skin tumors, thus being the leading cause of morbidity and mortality due to neoplasms. Its high mortality rate is associated with late diagnosis of the disease, when it is already in very advanced stages^1^.

Furthermore, Cervical Cancer (CC) is in the fourth position of the most frequent tumors among women worldwide^2^. This cancer represents a considerable indicator of inequality, since approximately 85% of its cases affect women with low education and low socioeconomic conditions^3^.

The SARS-CoV-2 virus, responsible for the COVID-19 pandemic, has brought negative impacts with regard to elective care in most countries, such as cancer screening, as these services have been interrupted to prioritize the reduction of the risk of spreading the new coronavirus in health services^4^.

Therefore, because it is a public health problem with global impact, high mortality rates, and a direct influence on the quality of life of the patient, it is important to encourage studies on this topic to try to understand the impacts that the pandemic had on early detection, screening, and diagnosis of cancer.

Thus, this study aims to evaluate the epidemiological behavior of hospital admissions for cervical and breast cancer in Brazilian women, as well as the performance of screening tests and diagnosis of these diseases in the years from 2010 to 2022 in Brazil.

## Materials and methods

The present study is an ecological time-series research, carried out using data obtained from secondary sources. It is a study that takes place in Brazil and its main focus is the epidemiological behavior of screening and diagnosis of breast and cervical cancer between 2010 and 2022. The main reason for the analysis of the last twelve years of hospitalizations for these neoplasms and their screenings is to identify how these variables have behaved and the influence that the pandemic of COVID-19 had on screening and early diagnosis.

The data were collected on October 26, 2022 from the national platform of Brazilian public health data called DATASUS, and the data for the present study were taken from four information systems: 1) Hospital Information System (SIH); 2) Cervical Cancer Information System (SISCOLO); 3) Breast Cancer Information System (SISMAMA) and 4) Cancer Information System (SISCAN), of public domain and managed by the Ministry of Health of Brazil. The purpose of these information systems is to compile data from hospitalization and exams offered to the population in the Brazilian Primary Care and Hospital Network. The data can be consulted on the site <https://datasus.saude.gov.br/informacoes-de-saude-tabnet/>.

The dependent variables of the study are: hospitalizations for cervical and breast cancer; number of cytopathological exams of the uterus in the Brazilian screening age group of 25 to 69 years; number of mammograms performed in the age group between 50 and 69 years. The independent variable used was the time period from January 1, 2010 to August 31, 2022, and Brazilian regions.

To achieve the objective of the study, an analysis was performed from the raw data, calculating the rate of hospitalization for cervical and breast cancer, separately, in addition to the rates of performance of cervical cytopathological exams and mammography by regions of Brazil. In order to do this, it was necessary to divide the number of hospitalizations (H) and exams by the number of the analyzed population, which are women of all ages to calculate the overall hospitalization, women in the age group 25 to 69 years for the cytopathological exam, and between 50 and 69 years for the mammography denominator, and finally multiplied by 100,000 for hospitalizations and multiplied by 1,000 in the case of exams, as shown in the equations below.

1. H = (Hospitalization for breast cancer/women of all ages) X 100,000
2. H = (Hospitalization for cervical cancer/women of all ages) X 100,000
3. H = (Mammograms/women aged 25-69) X 1,000
4. H = (Cervical cytopathology/women aged 50-69) X 1,000

After being collected, the data downloaded in CSV were processed and stored in Microsoft Excel®, where the database treatment occurred, selecting the important data for the research. The database was formatted to the standards of the JoinPoint® program, a software used in the statistical analysis of cancer morbidity and mortality, as it allows trends to be observed, whether stationary, ascending or descending, using Joinpoint models, guiding a historical series through Poisson regression to estimate Annual Percentage Change (APC) and Average Annual Percentage Change (AAPC) that can be consulted on the site <https://surveillance.cancer.gov/>. For each trend detected, a 5% significance level was used^5^.

The information obtained for the elaboration of the research came from secondary sources and were collected by public domain databases and, therefore, did not need to be submitted to the Research Ethics Committee appreciation, as recommended by the Brazilian Resolution No. 510, of April 07, 2016^6^.

## Results

The results obtained can be seen in Fig 1, which shows the rates of hospitalization for cervical and breast cancer. In Brazil, it is possible to observe a downward behavior in hospitalizations for cervical neoplasia between the years 2010 and 2015, which went from 47 hospitalizations per 100,00 women to 41 hospitalizations per 100,000 women. Then, it shows a growth until 2019, where it peaked with more than 48 hospitalizations per 100,000 women.

**Fig 1.**
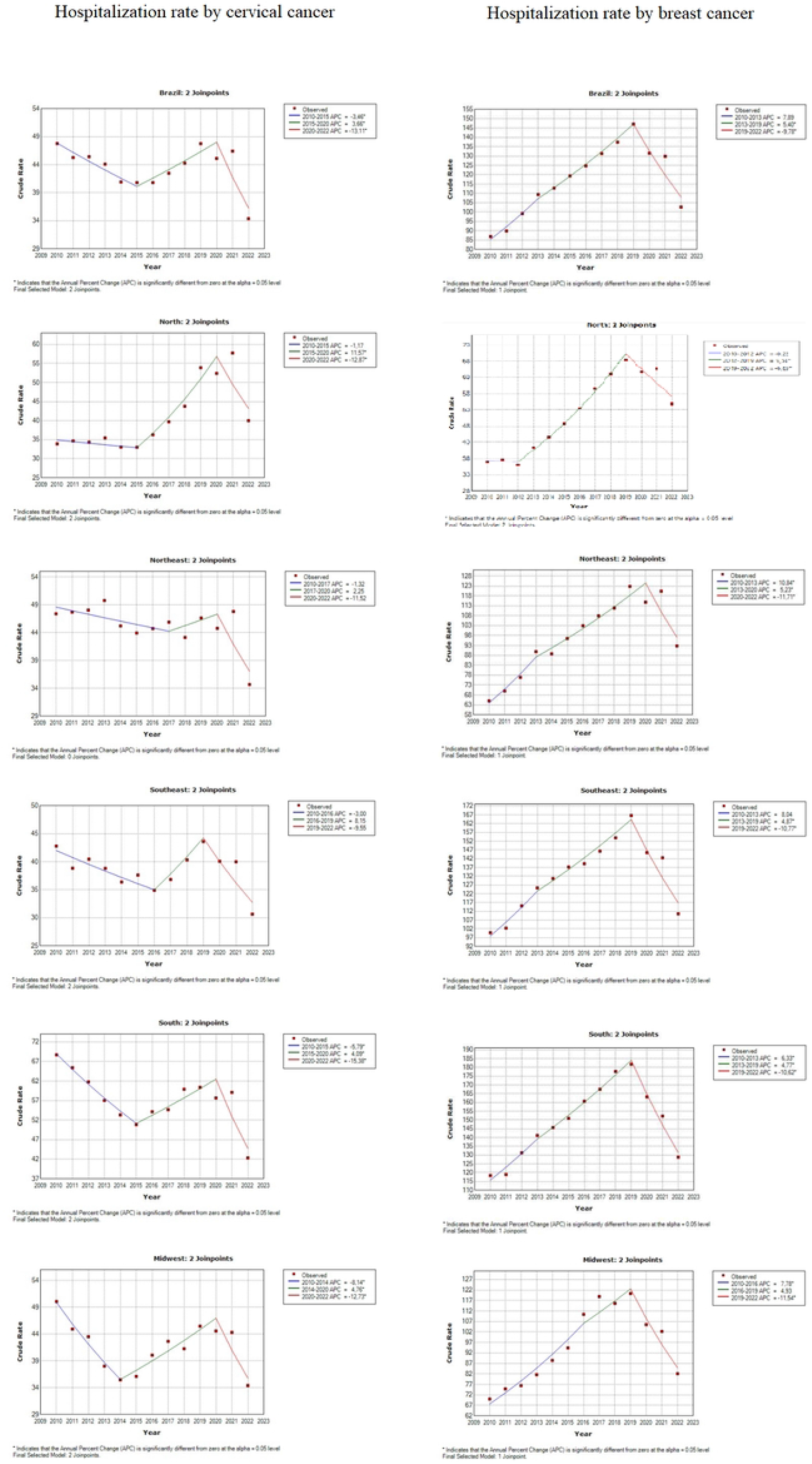
Hospitalization rate for cervical and breast cancer in Brazil, 2010 to 2022. Brazil, 2022.

The North region shows a more accentuated growth between 2015 and 2021, increasing from 33 to 58 hospitalizations per 100,000 women. The Northeast region shows more stability in hospitalization rates, varying between 43 and 50 hospitalizations per 100,000 women, with the exception of 2022, which showed 35 hospitalizations per 100,000 women. The Southeast, South and Midwest regions show a graph similar to Brazil, but with special emphasis on the South, which shows the highest hospitalization rates, ranging from 51 to 69 hospitalizations per 100,000 women, and reaching the lowest number in 2022, with 42 hospitalizations per 100,000 women.

In the pandemic years, between 2020 and 2022, the graph shows a downward trend in Brazil and in all regions, showing a reduction in hospitalizations. It is worth noting that the data for 2022 refer to the first eight months of the year, according to the collection date of the study.

The Table 1, which presents the statistical data of the linear regression of cervical cancer hospitalization in Brazil and their respective regions between 2010 and 2022, shows the presence of two Joinpoints in Brazil, in 2015 and 2020, the same occurs in the North, Southeast and South regions. The behavior changes in the Northeast regions, where the first Joinpoint is later, in 2017, and in the Midwest, which presents the earliest first Joinpoint, in 2014.

**Table 1.**
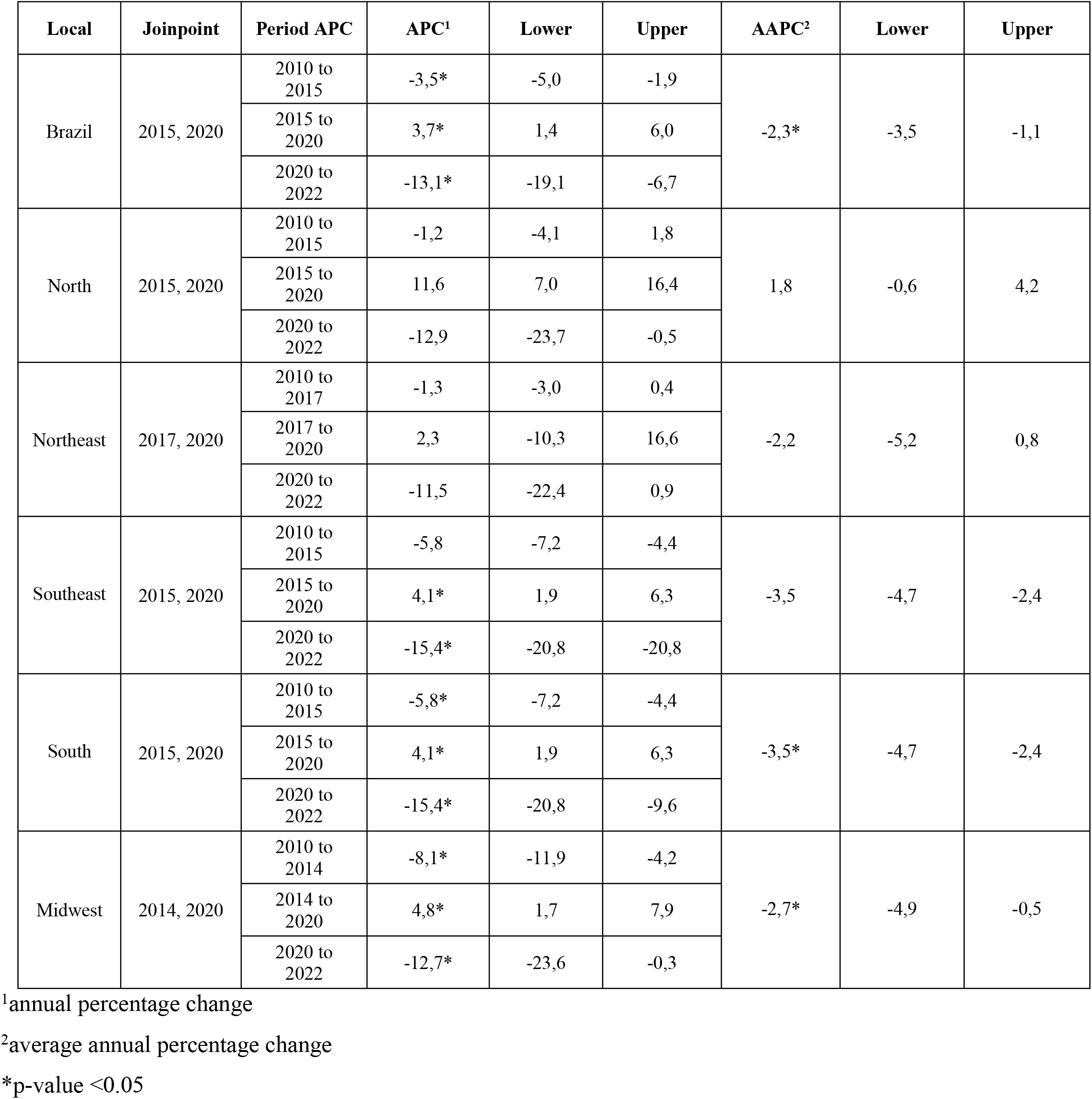
Joinpoint analysis of cervical cancer hospitalization rate in Brazil and its regions, 2010 to 2022. Brazil, 2022.

On the other hand, the hospitalization rates for breast cancer in Brazil and regions exceed the numbers for cervical cancer. In the national context, breast cancer shows a gradual growth behavior until 2019, peaking at 147 hospitalizations per 100,000 women. Southeast and South had higher peaks than Brazil, especially the latter, with 181 hospitalizations per 100,000 women. All regions grew to a peak in 2019 and then began to significantly regress the hospitalization rate, which shows a downward trend during the pandemic and may extend into the coming years.

Table 2, which presents the statistical data of the linear regression of breast cancer hospitalization in Brazil and their respective regions between 2010 and 2022, shows the presence of two Joinpoints in Brazil, in 2013 and 2019, the same occurs in the Southeast and South regions. On the other hand, in the North region, a regression of Joinpoint beginning in 2012 is perceived, the shortest observed. Being, therefore, a scenario similar to that of cervical cancer. The behavior changes in the Northeast, where the second Joinpoint is later, in 2020, and in the Midwest, where the first Joinpoint is later, in 2016.

**Table 2.** Joinpoint analysis of breast cancer hospitalization rate in Brazil and its regions, 2010 to 2022. Brazil, 2022.

| Local | Joinpoint | Period | APC<sup>1</sup> | Lower | Upper | AAPC<sup>2</sup> | Lower | Upper |
| --- | --- | --- | --- | --- | --- | --- | --- | --- |
| Brazil | 2013, 2019 | 2010 to 2013 | 7,9 | -0,1 | 16,5 | 2,0 | -0,5 | 4,5 |
|  |  | 2013 to 2019 | 5,4* | 1,8 | 9,1 |  |  |  |
|  |  | 2019 to 2022 | -9,8* | -16,5 | -2,5 |  |  |  |
| North | 2012, 2019 | 2010 to 2012 | -0,2 | -12,7 | 14,1 | 3,6* | 1,2 | 6,1 |
|  |  | 2012 to 2019 | 9,6* | 7,1 | 12,1 |  |  |  |
|  |  | 2019 to 2022 | -6,7* | -12,7 | -0,2 |  |  |  |
| Northeast | 2013, 2020 | 2010 to 2013 | 10,8 | 7,8 | 14,0 | 3,5* | 2,5 | 4,5 |
|  |  | 2013 to 2020 | 5,2* | 4,2 | 6,2 |  |  |  |
|  |  | 2020 to 2022 | -11,7* | -16,5 | -6,7 |  |  |  |
| Southeast | 2013, 2019 | 2010 to 2013 | 8,0 | -0,7 | 17,6 | 1,5 | -1,2 | 4,3 |
|  |  | 2013 to 2019 | 4,9* | 1,0 | 8,9 |  |  |  |
|  |  | 2019 to 2022 | -10,8* | -18,0 | -2,9 |  |  |  |
| South | 2013, 2019 | 2010 to 2013 | 6,3* | 1,5 | 11,4 | 1,1 | -0,4 | 2,6 |
|  |  | 2013 to 2019 | 4,8* | 2,6 | 7,0 |  |  |  |
|  |  | 2019 to 2022 | -10,6* | -14,7 | -6,4 |  |  |  |
| Midwest | 2016, 2019 | 2010 to 2016 | 7,8* | 5,3 | 10,3 | 1,9 | -1,2 | 5,1 |
|  |  | 2016 to 2019 | 4,9 | -8,5 | 20,3 |  |  |  |
|  |  | 2019 to 2022 | -11,5* | -17,4 | -5,3 |  |  |  |

<sup>1</sup>annual percentage change
<sup>2</sup>average annual percentage change
\*p-value <0.05

In Fig 2, it is shown the rate of cervical cytopathological exams and mammography performed from 2010 to 2022 in all regions of Brazil. Thus, it was possible to observe that in all charts of cytopathological tests of the uterine cervix, the behavior of the curve first goes through a drop, then the number of tests performed rises, and lastly, there is a reduction in recent years, with a more significant drop in the pandemic period, between 2020 and 2022.

**Fig 2.**
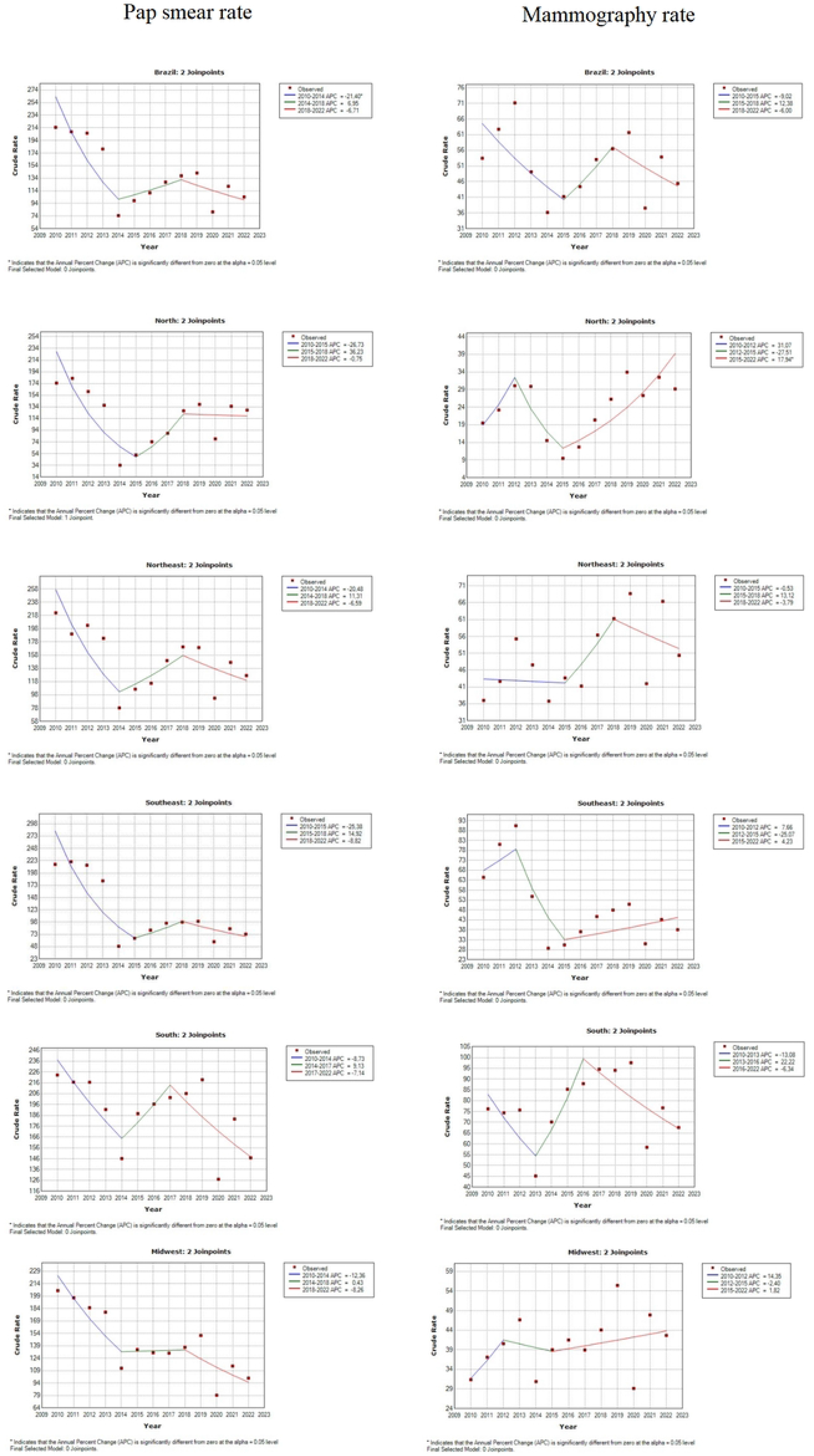
Pap smear and mammography rate in Brazil, 2010 to 2022. Brazil, 2022.

In Brazil, the rates vary between 78 and 218 exams per 1000 women, with special emphasis on the year 2014, which represents the worst index of the analyzed period, although there is growth afterwards, the numbers fall again starting in 2020. The same behavior is repeated in all regions of the country. The South is the region with rates that vary in a higher range, between 126 and 226 cytopathological exams per 1000 women. On the other hand, the North region presents the lowest statistics, between 34 and 182 exams per 1000 women.

Table 3, which presents the statistical data of the linear regression of the performance of cervical cytopathological exams in Brazil and their respective regions, between 2010 and 2022, shows the presence of two Joinpoint in Brazil, in 2014 and 2018, the same occurs in the Northeast and Midwest regions, differently from what was observed in hospitalizations. The behavior changes in the North and Southeast regions, where the first Joinpoint is later, in 2015, and in the South, which has the second earliest Joinpoint, in 2017.

**Table 3.**
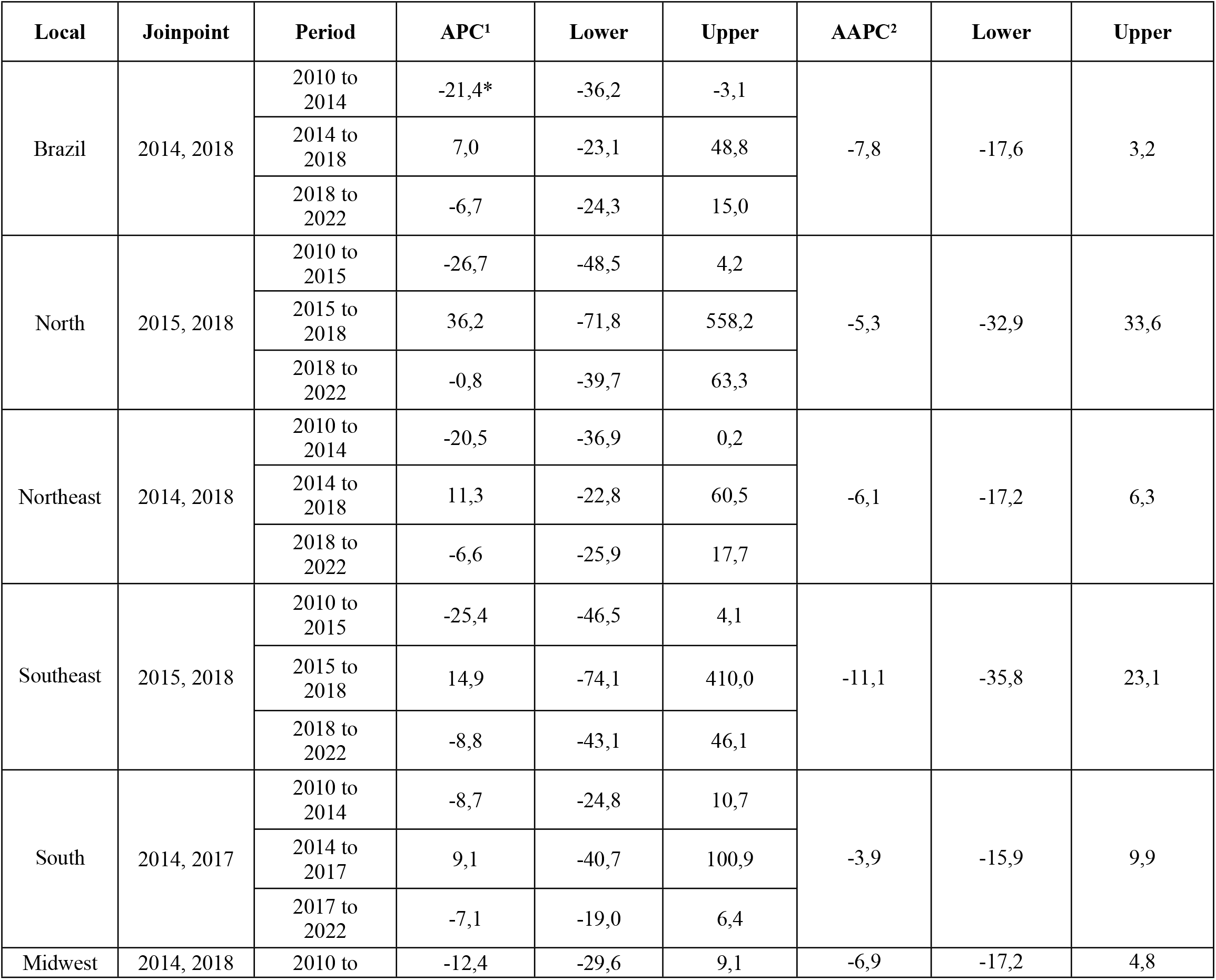
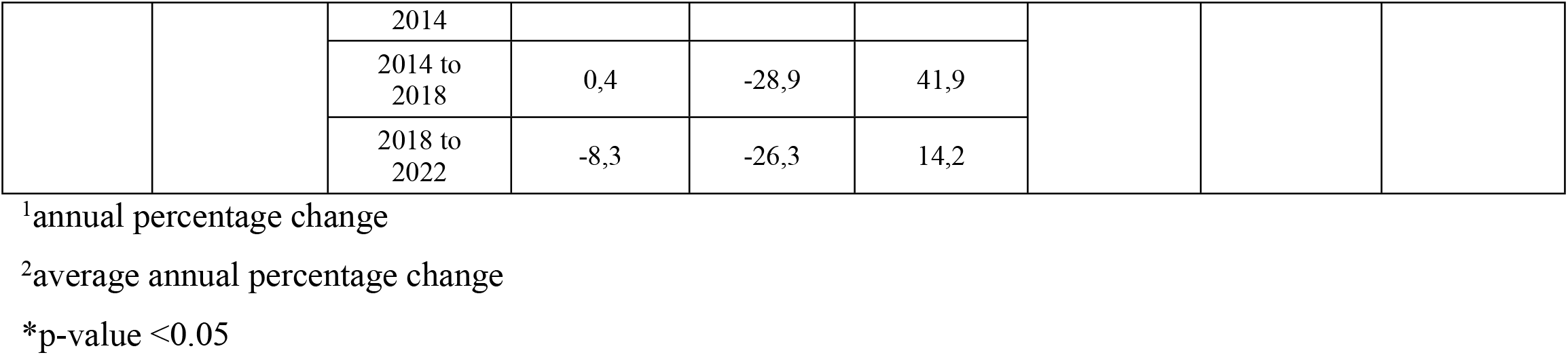
Joinpoint analysis of Pap smear rate in Brazil and its regions, 2010 to 2022. Brazil, 2022.

Regarding mammography exams, a unique behavior is observed in each region of the country. Nationally, the rates vary between 36 and 71 mammograms per 1000 women, with 2014 being the year with the lowest rate. However, it shows growth until 2019, falling again in 2020 and the following years, as do the Southeast and Midwest. The North, however, manages to increase the number of mammograms starting in 2015 and stabilize the number of the exam during the pandemic years. The Northeast and South regions show an upward trend, peaking in 2019, with 69 and 98 mammograms per 1000 women, respectively, but reduce the amount offered during the pandemic.

Furthermore, it is noteworthy that the Southern region stood out in the quantity of mammograms offered in the studied period, which varied between 45 and 98 exams per 1000 women. The North region, on the other hand, is a negative highlight in this aspect, since it presented rates that ranged between 9 and 34 mammograms per 1000 women.

Table 4, which presents the statistical data of the linear regression of the performance of mammography in Brazil and their respective regions between 2010 and 2022, shows the presence of two Joinpoints in Brazil, in 2015 and 2018, the same occurring in the Northeast regions. The behavior changes in the North, Southeast and Midwest regions, where the Joinpoint are earlier, being in 2012 and 2015, and in the South, which also presents earlier Joinpoints, in 2013 and 2016.

**Table 4.** Joinpoint analysis of mammography rate in Brazil and its regions, 2010 to 2022. Brazil, 2022.

| Local | Joinpoint | Period | APC<sup>1</sup> | Lower | Upper | AAPC<sup>2</sup> | Lower | Upper |
| --- | --- | --- | --- | --- | --- | --- | --- | --- |
| Brazil | 2015, 2018 | 2010 to 2015 | -9,0 | -25,1 | 10,4 | -3,0 | -19,8 | 17,2 |
|  |  | 2015 to 2018 | 12,4 | -52,8 | 167,5 |  |  |  |
|  |  | 2018 to 2022 | -6,0 | -28,5 | 23,7 |  |  |  |
| North | 2012, 2015 | 2010 to 2012 | 31,1 | -43,5 | 204,1 | 6,3 | -12,9 | 29,7 |
|  |  | 2012 to 2015 | -27,5 | -68,8 | 68,2 |  |  |  |
|  |  | 2015 to 2022 | 17,9* | 5,4 | 32,0 |  |  |  |
| Northeast | 2015, 2018 | 2010 to 2015 | -0,5 | -17,7 | 20,2 | 1,6 | -15,6 | 22,3 |
|  |  | 2015 to 2018 | 13,1 | -51,6 | 164,2 |  |  |  |
|  |  | 2018 to 2022 | -3,8 | -26,4 | 25,8 |  |  |  |
| Southeast | 2012, 2015 | 2010 to 2012 | 7,7 | -44,8 | 110,1 | -3,5 | -17,6 | 13,0 |
|  |  | 2012 to 2015 | -25,1 | -61,6 | 46,2 |  |  |  |
|  |  | 2015 to 2022 | 4,2 | -4,7 | 14,0 |  |  |  |
| South | 2013, 2016 | 2010 to 2013 | -13,1 | -33,7 | 13,9 | -1,7 | -12,9 | 10,8 |
|  |  | 2013 to 2016 | 22,2 | -28,8 | 109,9 |  |  |  |
|  |  | 2016 to 2022 | -6,3 | -14,5 | 2,6 |  |  |  |
| Midwest | 2012, 2015 | 2010 to 2012 | 14,3 | -48,1 | 151,8 | 2,7 | -14,8 | 23,8 |
|  |  | 2012 to 2015 | -2,4 | -55,7 | 114,9 |  |  |  |
|  |  | 2015 to 2022 | 1,8 | -8,4 | 13,1 |  |  |  |

<sup>1</sup>annual percentage change
<sup>2</sup>average annual percentage change
\*p-value <0.05

## Discussion

Brazil’s Unified Health System (SUS) is public and free for all Brazilian citizens and immigrants, and is complemented by the private health system, which also offers assistance through individually paid plans and procedures paid for by SUS to be offered free of charge to the population.

Health care in Brazil is divided into levels of complexity, with great focus on Primary Health Care (PHC) and the Hospital Network. In PHC, the Family Health Strategy (FHS) has been expanded, with changes in the financial and administrative structure, with a focus on participatory care for the community^7^. Thus, there are health units deployed in strategic locations with an adscribed population, aiming to achieve total coverage to ensure full, universal and free access to the entire population of the country^8^.

The Brazilian health model has similar characteristics to the models practiced in the United Kingdom and Canada, especially about the presence of a general doctor and a multidisciplinary team focused on family care^9^. The coverage of the FHS increased exponentially from the 2000’s, when 50.9% of the Brazilian population was covered, currently having an average coverage of 62.6%^10^. However, authors point out weaknesses that hinder the consolidation of this model of care, such as the difficulty in rapid access to tests and specialized consultations, a barrier that impairs the screening and early diagnosis that underlie the preventive characteristic and health promotion of PHC^11^.

Screening and early diagnosis are characterized as two strategies for the detection of cancer, which is a pathology that generates a high morbidity rate in Brazil, being a public health problem, especially when it affects priority groups, such as women, who have a high incidence of cervical and breast cancer, as shown in Fig. 1. About the screening, the objective is to discover pre-neoplastic lesions and pre-clinical cancer, through routine elective exams in a population without symptoms and/or signs suggestive of the cancer screened. Early diagnosis, on the other hand, seeks to recognize cancer at an early stage in a target population that presents suspicious symptoms of the disease^12^.

The early diagnosis allows higher chances of cure, better prognosis, and lower risk of hospitalization. This strategy is called “stage-shift”, when simpler and more effective therapies are used to minimize the stage of cancer presentation^13^. Three steps are important for the early diagnosis of cancer, which are awareness and search for health care, clinical and diagnostic evaluation, and lastly, access to treatment. The impact of the early diagnosis of cancer permeates both an improvement in the patient’s quality of life and a reduction in hospitalization costs^12^.

Cervical Cancer (CC) is the third most prevalent type of cancer among women in Brazil, with HPV (Human Papilloma Virus) as the main cause. According to Brazilian National Cancer Institute (INCA) estimates, for 2022, about 16,590 women will be affected in new cases, which represents a risk considered to be 15.38 cases per 100,000 women^14^. Despite the high prevalence, the screening services offered by SUS, despite being opportune, show a decrease in the incidence of this type of cancer, as well as the mortality caused by it^15^.

France, as well as other countries in Europe, recommends the use of HPV testing as a primary screening tool for cervical cancer in July 2019^16^. One year later, these indications were officially included in the national CC screening program through a ministerial order (Ministère des Solidarités et de la santé, 2020) as well as a reference for program evaluation comprising performance indicators was developed^17^.

The transition to fully effective primary HPV screening can be time-consuming and complicated. The Netherlands, for example, was the first country in Europe to implement primary HPV screening in 2017. The new program started only after a preparatory phase that lasted more than 4 years^17^.

For the diagnosis and screening of CC, in Brazil, the main test used in SUS is the Pap smear or cytopathological examination of the uterine cervix, guaranteed to all people with a uterus, in the age group of 25 to 64 years, who have already started sexual life, throughout the national territory. This age group is a priority because it has a higher incidence of high-grade lesions that are precursors of cervical cancer^8^.

However, difficulties are still faced for the full offer of this procedure, as an example, there is the exit, in 2008, of the Pap smear from the Fund for Strategic Actions and Compensation (FAEC), which is a public budget intended exclusively for strategic actions. In this funding model, the procedure was performed and registered in the system for direct receipt of funds^18^.

Since 2008, Pap smears have been funded by the Medium and High Complexity Funding (MAC), which transfers the financial resources to state and municipal managers to distribute among all the procedures performed in the service^19^. This action may have contributed significantly to the reduction in the performance of cervical cytopathological exams nationwide between 2010 and 2014, as shown in Fig 2.

Another difficulty encountered is the periodicity in which the Pap smear is performed, since the Ministry of Health recommends that it should be done every three years when the woman has a healthy cervix. However, data show that only 8% of the exams are performed every three years, while there are about 50% of annual repetitions. These data show that some women do not perform the exam regularly, while others repeat it excessively^19^.

According to Fig 2, it is possible to observe that from 2014 on, there is an increase in the rate of cervical cytopathological exams, an occurrence that may be linked to the new Brazilian norm through Ordinance no. 3.388, of December 30, 2013, which redefines the National Qualification in Cytopathology in the prevention of cervical cancer (QualiCito), aiming to increase the coverage of cytopathological exams, collect samples with a higher level of quality, devise means of improving professionals, in addition to applying an incisive monitoring through SISCAN^20^.

Breast carcinoma is a heterogeneous disease and comprehends biologically distinct neoplasms, with varied clinical and morphological symptoms. Still, there are more hostile subtypes, which transition to the form of metastasis in adjacent or distant organs, while most tumors have less aggressive characteristics and, consequently, a better prognosis^18^.

Thus, mammography is the gold standard test for early diagnosis and screening of this neoplasm because, although there are limitations, this procedure is still the most efficient to detect masses, lesions or non-palpable nodules. The recommended age range for mammography is 50 to 69 years, once every two years^18^.

This orientation is implemented in most countries that have adopted organized screening for breast neoplasia, due to evidence proving an effective impact on mortality reduction and improved risk-benefit, which is not noted in other age groups^21,22^.

However, it is observed a divergence of situations related to the provision of mammography equipment in Brazil, covering several adversities in the organization of the service, such as scarcity of available equipment, trained professionals, obstacles of access related to geographic distance and underutilization of equipment^23^. This problem is reflected in the results of the present study (Fig 2), in such a way that the South and Southeastern regions, with greater health structures and greater financial resources, present higher rates of mammography, demonstrating the heterogeneity in the public health in the country.

Worldwide, breast cancer is the most common cancer among women. So that the highest incidence rates found in studies were in Australia and New Zealand, the countries of Northern Europe and Western Europe. Research shows that regardless of the socioeconomic factors in the country, new cases of breast cancer are in the top positions of malignant neoplasms in women^24,25^.

Nevertheless, in developed countries, a decrease in the incidence of breast neoplasms has been detected, which may be associated with the decrease in hormone replacement treatment in women in the climacteric period^24,25^, but this behavior is different from what is observed in Brazil, because, as shown in Fig 1, between 2010 and 2019, hospitalizations for breast cancer follow a gradual increase in hospitalization.

A factor of great impact in recent years has been the pandemic caused by COVID-19, which has reflected on the performance of examinations and surgical procedures worldwide^26^. The treatment of breast carcinoma has undergone considerable delays and cancellations, mainly because it is an area that needs multi-professional cooperation^27^. The results of the authors Demarchi et al. (2022)^28^, who analyzed a decrease of 1,705,475 mammograms in Brazil in the year 2020 alone, show a reduction of about 40% in relation to the previous year.

Nonetheless, according to the results obtained in this study, some regions, such as the North, Midwest and Southeast, showed an upward behavior in the performance of mammograms. This finding may be related to the increase in financial resources received by the states and municipalities to invest in the health system during the pandemic period. In addition, it is known that the new financing model - MAC - allows the manager to apply the resources for each procedure independently, which may cause a heterogeneity in the supply of mammograms among the regions^29^.

As a result of the fast spread of the virus and the recommendations for social distancing promoted by the WHO to decrease cases of COVID-19, the Society of Breast Imaging proposed delaying mammograms by “several weeks or a few months”^30^. In contrast, the Canadian Society of Breast Imaging and the Canadian Association of Radiologists proposed that all mammograms should be delayed for at least 6 to 8 weeks^31^. However, in the country, the Brazilian Society of Mastology (SBM) issued in March a statement attesting that the conduct should be determined according to the local reality, taking into consideration the risks and demands posed by the pandemic^32^, which corroborates the different rates of mammography among the regions, as demonstrated in Fig 2.

Regarding the average number of Pap smears performed in the first period of the pandemic, it is observable from the results of a study conducted in Ontario, Canada, an 85.8% drop in the number of Pap smears performed in the first period of the pandemic in its worst month, with an overall drop of 63.8% between the month of March and August 2020^33^.

Another study, conducted in the United States, California, points to a 78% reduction in the monthly rate of Pap smears performed by women aged 21 to 29 years and 82% in women aged 30 to 65 years^34^. This pattern is not very different from that observed in the present study, in which Brazil, showed a drop from 134 preventive exams per thousand women to approximately 95 exams from 2018 to 2022, respectively, reflecting the impact of the pandemic period on the performance of these exams.

Lastly, the limitations of this study were related to the availability of data regarding the quantification of materials made available for the performance of mammography and Pap smears nationwide, as well as more precise information regarding the financing of these materials.

## Conclusion

Knowing that breast cancer and cervical cancer occupy significant positions in female morbidity and mortality in Brazil, it is prudent to point out suitable solutions to combat these diseases. Thus, the screening methods must be a priority, with the standardization of the wide attendance and availability of Pap smears and mammograms throughout the country.

However, currently there are impasses for the full offer of these procedures, even with the various public policies for the promotion of this field. In addition, it was observed in the study that hospitalization rates for these neoplasms had a gradual increase before the pandemic and that, after 2020, there was a reduction both in hospitalization and in the supply of diagnostic tests. Therefore, there is a need for further studies that highlight the impact of the pandemic period on the detection, screening, and early diagnosis of these diseases.

## Data Availability

All relevant data are within the manuscript.

## Conflict of Interest

The authors declare that no competing interests exist regarding this study.

## Financing

This study will be partially funded by the Coordination for the Improvement of Higher Education Personnel-Brazil (CAPES): Financial Code 001. Funders will have no role in the study design, data collection and analysis, publication decision or preparation of the manuscript.

## Notes

### Competing Interest Statement

The authors have declared no competing interest.

### Funding Statement

GNC, TNBM, KTSC are funded by the Coordination for the Improvement of Higher Education Personnel- Brazil (CAPES): Financial Code 001. Funders will have no role in the study design, data collection and analysis, publication decision or preparation of the manuscript.

### Author Declarations

The information obtained for the elaboration of the research came from secondary sources and were collected by public domain databases and, therefore, did not need to be submitted to the Research Ethics Committee appreciation, as recommended by the Brazilian Resolution No. 510, of April 07, 2016.

